# Novel biomarkers for distinguishing bacterial from non-bacterial infection: a systematic review

**DOI:** 10.1101/2025.09.03.25334997

**Authors:** Holly Drummond, Clare Mills, Helen Groves, Tom Waterfield

## Abstract

**Background:** Accurately distinguishing bacterial from non-bacterial infections is essential to guide antibiotic use but remains clinically challenging. Current biomarkers such as C-reactive protein and procalcitonin have limited diagnostic accuracy.

**Objective:** To identify novel blood protein biomarkers with clinically acceptable performance for differentiating bacterial from non-bacterial infections.

**Methods:** Following PRISMA guidelines, we systematically searched Medline and prior reviews (2010–2024) for studies evaluating the diagnostic accuracy of protein biomarkers in serum or plasma for bacterial infection. Biomarkers were considered clinically useful if sensitivity ≥90%, specificity ≥80%, or AUC ≥0.9, was reported in studies with low risk of bias (assessed via modified QUADAS-2).

**Results:** From 2,236 records, 47 studies were included, assessing 50 individual biomarkers and 12 signatures. Two individual biomarkers (LCN2, IFN-α) and three multi-marker signatures (TRAIL+IP-10+CRP, E-selectin+IL-18+NCAM1+LCN2+IFN-γ, and E-selectin+IL-18+NCAM1+LG3BP+LCN2+IFN-γ) met pre-defined performance thresholds. TRAIL+IP-10+CRP showed consistent performance across six studies.

**Conclusions:** TRAIL+IP-10+CRP and LCN2 are promising host-response biomarkers for bacterial infection. Multi-marker panels may enhance diagnostic accuracy but require further validation. These tools have the potential to improve clinical decision-making and reduce unnecessary antibiotic use.

## Introduction

Infectious diseases remain a leading cause of global morbidity and mortality, accounting for a significant proportion of the worldwide disease burden (Vos et al., 2020). Despite advances in prevention and treatment, the accurate and timely diagnosis of bacterial infections remains a persistent challenge. Differentiating bacterial from non-bacterial infections, such as viral or inflammatory conditions, is often clinically difficult, as signs and symptoms may overlap (Van den Bruel & Thompson, 2014; van Houten et al., 2017). This diagnostic uncertainty leads to the empirical use of antibiotics, contributing to unnecessary antimicrobial exposure, prolonged hospital stays, increased healthcare costs, and the growing problem of antimicrobial resistance (AMR) (Laxminarayan et al., 2013; Ventola, 2015).

The current gold standard for diagnosing bacterial infection, microbial culture from sterile sites, is limited by long turnaround times, risk of false negatives due to prior antibiotic use, and sample contamination (Iroh Tam et al., 2015). Molecular methods like real-time PCR offer faster results and improved sensitivity in some contexts, but still face challenges, including limited pathogen panels and an inability to distinguish colonisation from active infection (Shah et al., 2023).

Given these limitations, there is increasing interest in host-biomarker-based diagnostics. By measuring the host response to pathogens, clinicians may better distinguish bacterial from non-bacterial causes of illness (Habgood-Coote et al., 2023; van Houten et al., 2017). Host biomarkers like C-reactive protein (CRP) and procalcitonin (PCT) are widely used but have shown suboptimal sensitivity and specificity, especially in early disease (Norman-Bruce et al., 2024). Emerging host protein biomarkers and multi-marker protein signatures report improved diagnostic accuracy and are well-suited for integration into clinical platforms (Gunaratnam et al., 2021; Jackson et al., 2023; Leticia Fernandez-Carballo et al., 2021).

Previous systematic reviews have evaluated protein biomarkers of bacterial infection, however, the rapidly growing body of evidence, which includes newly identified markers and validation studies of previously reported candidates, requires an updated synthesis. This systematic review aims to provide an up-to-date evaluation of novel protein biomarkers and evaluate which report clinically acceptable diagnostic performance for the differentiation of bacterial infections.

## Methods

This systematic review was conducted in accordance with the Preferred Reporting Items for Systematic Reviews and Meta-Analyses (PRISMA) guidelines (Page et al., 2021).

### Eligibility criteria

Studies were eligible if they assessed the diagnostic performance of novel blood protein biomarkers measured in plasma or serum for distinguishing bacterial from non-bacterial infections. Studies evaluating novel biomarkers alongside established markers such as PCT or CRP were included if data on the novel biomarkers alone could be extracted. Exclusion criteria included studies conducted exclusively in immunocompromised populations, laboratory models, or animal models.

### Information sources

The primary literature search was conducted in Medline Ovid on January 11, 2024, covering publications from January 2019 to January 2024. Reference lists of select studies were also used as information sources. Additionally, studies included in two prior systematic reviews covering the period from January 2010 to May 2019 were compiled for screening (Kapasi et al., 2016; Tan et al., 2023).

### Search strategy

The search strategy was designed to identify studies reporting novel blood protein biomarkers capable of differentiating bacterial from non-bacterial infections. The full systematic literature search terms were: (((Biomarkers/ or Proteomics/ or (((biological or blood or serolog* or serum or infection* or Inflammat*) adj6 marker*) or biomarker* or proteomic* or proteogenomic*).ab,ti. or marker*.ti.) and (Bacterial Infections/ or Bacteremia/ or Meningitis, Bacterial/ or Pneumonia, Bacterial/ or Sepsis/ or Pneumonia/ or Meningitis/ or Urinary Tract Infections/ or ((bacterial* adj3 (infect* or superinfect*)) or bacteraemi* or bacteremi* or sepsis or septic or pneumoni* or meningitis or (urinary-tract adj3 infection*)).ab,ti.) and (exp Virus Diseases/ or Diagnosis, Differential/ or *Diagnosis/ or (((virus* or viral*) adj3 infection*) or ((different* or accura*) adj6 diagnos*)).ab,ti. or (sever* or serious or discriminat* or (bacter* adj3 viral) or predict* or diagnos*).ti.) and (exp Blood/ or Blood.fs. or (blood or serum or plasma or circulat*).ab,ti.)) not (exp animals/ not humans/)) and english.la. and (“2019” or “2020” or “2021” or “2022” or “2023” or “2024”).yr.

### Selection process

Screening and selection were performed independently by two reviewers (HD and CM) using the Rayyan online platform (Ouzzani et al., 2016). Titles and abstracts were initially screened, followed by full-text review of potentially eligible studies. Discrepancies were resolved through discussion.

### Data collection process

Data extraction was conducted independently by both reviewers using a standardised and piloted data extraction tool. Extracted data included biomarker type, diagnostic performance metrics, study population characteristics, and reference standards used.

### Data items

Key data items included sensitivity, specificity, and area under the receiver operating characteristic curve (AUC) for each biomarker. Studies were also assessed for sample type (plasma or serum), biomarker measurement platform and population characteristics.

### Study risk of bias assessment

Risk of bias and applicability were assessed using a modified version of the QUADAS-2 tool (Whiting et al., 2011). Studies were classified as low risk of bias if all four domains scored low. If any domain was rated as high or unclear, the study was considered at risk of bias.

### Effect measures

The reference standard was defined as the detection of a bacterial pathogen in blood, cerebrospinal fluid, urine, bronchoalveolar lavage fluid, sputum, or stool via culture or quantitative PCR. Studies relying solely on clinical diagnosis of sepsis without pathogen isolation were excluded.

### Synthesis methods

Study and biomarker information were displayed as a table. Biomarkers were considered clinically useful if they achieved sensitivity ≥90% and specificity ≥80%, or AUC ≥0.9, and were reported in studies with a low risk of bias, as based on established criteria for diagnostic performance (Dittrich et al., 2016).

### Reporting bias and Certainty assessment

It is acknowledged that studies reporting underperforming biomarkers or negative findings are less likely to be published, introducing a risk of reporting bias. This may lead to an overestimation of biomarker performance across the included studies. However, as the objective of this review was to identify biomarkers with excellent reported diagnostic characteristics for further evaluation, assessing publication bias was not considered informative for candidate selection. As described under bias assessment, the QUADAS-2 tool was used to assess the risk of bias and applicability concerns for each included diagnostic accuracy study. Only studies with low bias were considered for the evaluation of best-performing biomarkers to ensure high certainty of evidence.

## Results

### Study selection

A total of 2,236 studies were identified through the literature search. After screening titles, abstracts, and full texts, 47 studies met the inclusion criteria and were included in the final analysis (Figure 1).

**Figure 1:**
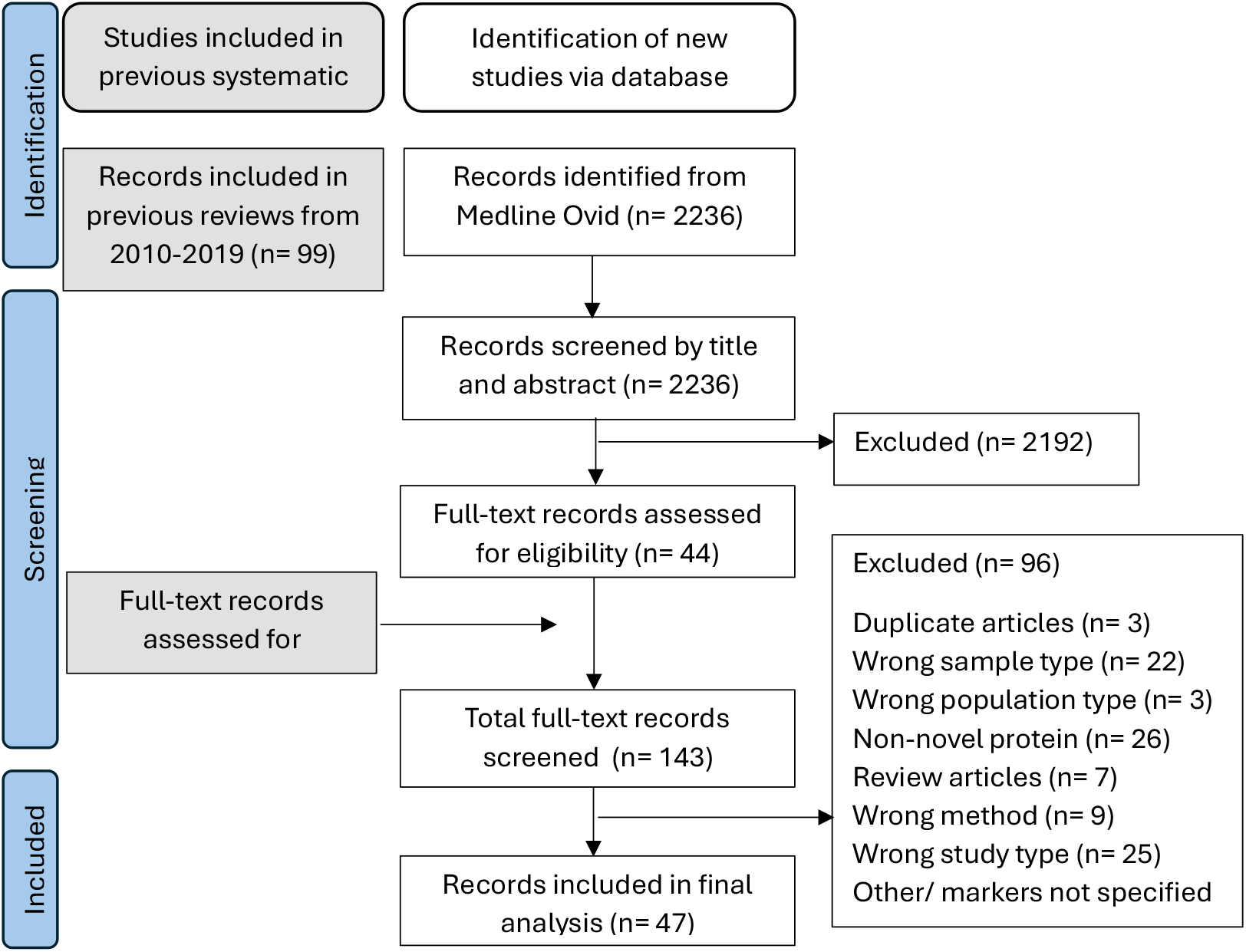
Flow diagram visualising the identification of publications, reasons for study exclusions and selection of studies for systematic review.

### Study Characteristics

A summary of the 47 included studies is presented in Supplementary Table 1. Most studies (79%, 37/47) were published between 2015 and 2023. Nearly half (45%, 21/47) focused exclusively on paediatric populations (≤18 years), 42% (20/47) on adults (≥18 years), 11% (5/47) included both, and 2% (1/47) did not report age ranges.

Across the studies, 50 individual biomarkers and 12 biomarker signatures were evaluated. The majority (72%, 45/62) were assessed in only one publication. The most commonly studied were LCN2 (19%, 9/47), TRAIL+IP-10+CRP (17%, 8/47), and IL-6 (15%, 7/47). The TRAIL+IP-10+CRP signature was evaluated in 2,638 participants, LCN2 in 2,220, and presepsin in 1,916.

### Risk of bias in studies

Methodological quality, assessed using the modified QUADAS-2 tool (Supplemental Table 2; Figure 2), was generally high. In the patient selection domain, 11% of studies had high risk of bias, while 83% were low risk and 6% unclear; applicability concerns were low in 92% of studies. In the index test domain, 98% of studies were low risk with no applicability concerns. For the reference standard, 15% showed high risk of bias, and 13% had high applicability concerns. The flow and timing domain showed the highest risk, with 36% of studies rated high risk of bias.

**Figure 2.**
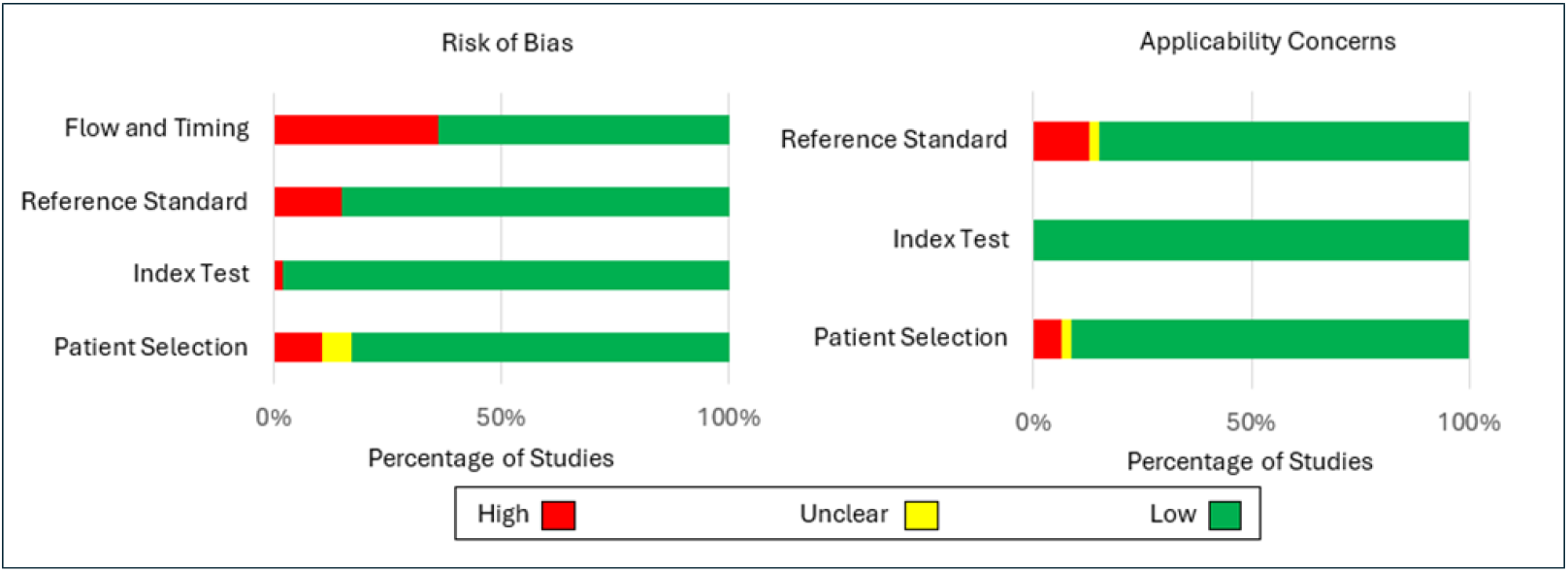
Graphical presentation of the Quality Assessment of Diagnostic Accuracy Studies-2 (QUADAS-2) assessment.

### Results of individual studies/biomarkers and syntheses

The literature search identified two individual biomarkers and three multi-marker signatures that met the pre-specified diagnostic performance thresholds for adequate clinical use (Table 1). The individual biomarkers were IFN-α and LCN2, while the grouped signatures included TRAIL+IP-10+CRP, a six-marker panel comprising E-selectin, IL-18, NCAM1, LG3BP, LCN2, and IFN-γ and a five-marker panel comprising E-selectin, IL-18, NCAM1, LCN2, and IFN-γ.

**Table 1.** Summary of the biomarkers and signatures that met the pre-specified diagnostic performance thresholds.

| Biomarker/<br>Signature | Number of<br>Publications | Reference | Sensitivity/<br>% | Specificity/<br>% | Area<br>under<br>the<br>curve |
| --- | --- | --- | --- | --- | --- |
| IFN-α | 1 | Trouillet-Assant et al., 2020 | | | 0.93 |
| LCN2 | 2 | Yu et al, 2016 | 98.5 | 94.3 | 0.97 |
|  |  | Wang et al, 2023 | 91.8 | 81.2 | 0.91 |
| TRAIL+IP-10+CRP | 6 | Ashkenazi-Hoffnung et al. 2018 | 93.5 | 94.3 |  |
|  |  | Srugo et al., 2017 | 93.8 | 89.8 |  |
|  |  | Chokkalla et al., 2023 | 94.0 | 88.0 |  |
|  |  | Halabi et al., 2023 | 98.1 | 88.4 |  |
|  |  | Oved et al., 2015 |  |  | 0.94 |
|  |  | van Houten et al., 2017 |  |  | 0.90 |
| E-selectin+IL18+<br>NCAM1+<br>LG3BP+LCN2+IFN-γ | 1 | Jackson et al, 2023 | 90.4 | 89.6 | 0.89 |
| E-selectin+IL18+<br>NCAM1+ LCN2+IFN-γ | 1 | | 93.1 | 80.4 | 0.89 |

IFN-α was evaluated in a single study (n=101) and showed excellent performance (AUC 0.93; (Trouillet-Assant et al., 2020). However, it should be noted that a Simoa®, a high-sensitivity quantification method, was used, which may not be compatible with available clinical platforms.

LCN2 met the pre-specified thresholds in two out of the nine relevant publications with AUCs of 0.97 and 0.91 (Wang et al., 2023; Yu et al., 2016).

The TRAIL+IP-10+CRP signature met the pre-specified thresholds in six publications. In two studies, the signature had AUCs ≥0.9 (0.94 and 0.90) (Oved et al., 2015; van Houten et al., 2017). The TRAIL+IP-10+CRP signature met the pre-specified thresholds in the remaining four papers with sensitivity of 93.5% and specificity of 94.3% (Ashkenazi-Hoffnung et al., 2018), sensitivity of 93.8% and specificity of 89.8% (Srugo et al., 2017), sensitivity of 94.0% and specificity of 88.0% (Chokkalla et al., 2023) and sensitivity of 98.1% and specificity of 88.4% (Hainrichson et al., 2023).

The E-selectin+IL18+NCAM1+LG3BP+LCN2+IFN-γ and E-selectin+IL18+NCAM1+LCN2 +IFN-γ signatures were studied in a single publication that comprised 306 subjects and met the pre-specified thresholds with a sensitivity of 90.4% and a specificity of 89.6% (Jackson et al., 2023).

## Discussion

This systematic review identified several promising blood protein biomarkers and multi-marker signatures for distinguishing bacterial from non-bacterial infections. Among 47 included studies evaluating 50 individual biomarkers and 12 multi-marker signatures, five candidates met pre-specified diagnostic performance thresholds for potential clinical utility: two individual biomarkers (IFN-α and LCN2) and three grouped signatures (TRAIL+IP-10+CRP, E-selectin, IL-18, NCAM1, LCN2, and IFN-γ and E-selectin+IL-18+NCAM1+LG3BP+LCN2+IFN-γ).

The most robust evidence was found for the TRAIL+IP-10+CRP signature, which demonstrated high sensitivity and specificity in six independent studies across diverse populations. This consistency in diagnostic performance and broad validation highlights its potential as a clinical tool for infection differentiation. This signature is available on a commercial platform from MeMed, and a recent meta-analysis of the signature has further highlighted its strong diagnostic performance (Kandula & Farrell, 2023).

LCN2, while frequently studied, showed variable performance, meeting diagnostic thresholds in only two of nine studies. The five and six-protein panels described were evaluated in a single large study but achieved high diagnostic accuracy, suggesting potential for further validation (Jackson et al., 2023). IFN-α also demonstrated excellent diagnostic performance (AUC 0.93) in a single study, but its reliance on ultra-sensitive Simoa® technology raises concerns regarding near-term clinical applicability, as more widely available immunoassay platforms (e.g., ELISA, Lateral flow) may not support this level of sensitivity (Trouillet-Assant et al., 2020).

Our findings align with earlier reviews (Kapasi et al., 2016; Tan et al., 2023), which also identified LCN2 and TRAIL-based markers as promising candidates. However, the addition of newly published data, including multi-marker signatures and expanded cohorts, strengthens the case for prioritising certain biomarkers for clinical translation. This review updates the field by synthesising evidence on novel biomarker combinations and highlights which biomarkers and combinations show adequate clinical performance.

This review has several limitations. First, heterogeneity in study design, populations, reference standards, and biomarker platforms precluded formal meta-analysis. Second, potential publication bias cannot be excluded; studies with negative or inconclusive findings are less likely to be published, possibly leading to an overestimation of diagnostic performance. Although QUADAS-2 was used to assess risk of bias, the overall certainty of the evidence may still be influenced by selective reporting.

Further validation of promising biomarkers, particularly the multi-marker signatures, in prospective, multicentre studies is essential. These studies should include well-characterised populations across all age groups and should use consistent reference standards for pathogen detection. For the biomarker combination CRP, TRAIL and IP-10, which has extensive validation, studies should focus on head-to-head comparisons with established markers (e.g., CRP, PCT) and integration into clinical decision-making tools.

## Conclusions

This systematic review identifies TRAIL+IP-10+CRP and LCN2 as the most promising biomarkers for distinguishing bacterial from non-bacterial infections, with strong supporting evidence across multiple studies. Emerging multi-marker signatures may further enhance diagnostic performance, though additional validation is needed. Incorporating host-response biomarkers into clinical practice has the potential to improve diagnostic accuracy, reduce unnecessary antibiotic use, and support global efforts to combat antimicrobial resistance.

## Data Availability

All data produced in the present study are available upon reasonable request to the authors

## Author Contributions

HD, CM, HG, and TW conceptualised and designed the study. HD and CM undertook the analysis. HD wrote the first draft of the report. All authors reviewed and approved the final manuscript and had full access to all the data.

## Registration

This review has not been registered

## Data availability

Full study data extraction and screening lists are available upon reasonable request.

## AI Use Statement

Artificial intelligence (ChatGPT by OpenAI) was used to assist with improving the clarity, grammar, and readability of the manuscript. The authors reviewed and edited all AI-generated content to ensure accuracy and integrity. No AI tools were used for data analysis or interpretation.

