## Supplemental material for "Novel biomarkers for distinguishing bacterial from non-bacterial infection: a systematic review"

**Supplemental Table 1.** Summary of study characteristics of the 47 included studies meeting eligibility criteria.

| Publication (reference) | Study setting | Study population | Age range | Study groups | Sample Size | Biomarker(s) | Sensitivity (%) | Specificity (%) | Area under curve |
|---|---|---|---|---|---|---|---|---|---|
| Chalupa *et al.* (2011) | Infectious diseases department, Czech Republic | Presenting with fever | 18-80y | bacterial vs viral | 81 | Interferon-γ | | | 0.58 |
| | | | | | | Heparin-binding protein | | | 0.83 |
| Conroy *et al.* (2014) | Outpatient unit, Colombia | Acute febrile syndrome less than 96 hours | 5-81y | bacterial vs viral | 175 | Angiopoietin-like 3 | 84.1 | 63.8 | 0.81 |
| | | | | | | Interleukin-18 binding protein | 94.7 | 83.0 | 0.92 |
| | | | | | | Interferon gamma-induced protein-10 | 82.3 | 72.3 | 0.84 |
| | | | | | | Platelet Factor 4 a | 39.8 | 97.9 | 0.69 |
| | | | | | | Soluble intercellular adhesion molecule-1 | 83.2 | 78.7 | 0.84 |
| | | | | | | Factor D | 69.0 | 93.6 | 0.85 |
| | | | | | | Soluble Endoglin | 79.7 | 93.6 | 0.92 |
| | | | | | | Soluble vascular endothelial growth factor-2 | 84.1 | 51.1 | 0.71 |
| | | | | | | Soluble angiopoietin-2 | 72.6 | 68.1 | 0.73 |
| Elsing *et al.* (2011) | General practitioner and emergency department, Germany | Suspected acute gastroenteritis | >18y | bacterial vs viral | 88 | Lipopolysaccharide binding protein | 82.0 | 67.0 | 0.76 |
| Haran, Buglione-Corbett and Lu (2013) | Emergency department, United States of America | Influenza-like illness symptoms, specifically fever and cough | >18y | bacterial vs viral | 80 | Interleukin-4 | 100 | 76.5 | 0.89 |
| | | | | | | Interleukin-5 | 85.7 | 67.6 | 0.82 |
| | | | | | | Interleukin-6 | 50.0 | 97.1 | 0.58 |
| | | | | | | Granulocyte-macrophage colony-stimulating factor | 78.6 | 80.9 | 0.85 |
| | | | | | | Interferon-γ | 100 | 88.2 | 0.94 |
| Katoh *et al.* (2014) | Inpatient unit, Japan | Influenza or pneumococcal pneumonia | >18y | bacterial vs viral | 83 | Galectin-9 | 81.4 | 75.0 | 0.85 |
| Mansour *et al.* (2011) | Paediatric emergency department, United States of America | Fever | 2weeks -14y | bacterial vs viral | 76 | Secretory phospholipase A2 | 93.0 | 67.0 | 0.89 |

| | | | | | | | | | |
|---|---|---|---|---|---|---|---|---|---|
| Siahanidou *et al.* (2014) | Intensive care unit, Greece | Neonates with infection | 4- 28 days | bacterial vs viral | 47 | Serum urokinase-type plasminogen activator receptor | | | 0.51 |
| te Witt *et al.* (2012) | Stored samples taken on admission, Netherlands | Ill-returned febrile travellers | | bacterial vs viral | 69 | Neopterin | 56.0 | 26.0 | 0.33 |
| ten Oever *et al.* (2012) | Emergency department, Netherlands | Suspected infection, ≥2 signs of sepsis | ≥16y | bacterial vs viral | 56 | Lipopolysaccharide binding protein+C-reactive protein | | | 0.85 |
| | | | | | | Interleukin-6+C-reactive protein | | | 0.84 |
| | | | | | | Soluble triggering receptor expressed on myeloid cell-1+C-reactive protein | | | 0.83 |
| | | | | | | Interleukin-18+C-reactive protein | | | 0.82 |
| Weh *et al.* (2013) | Infectious ward, Germany | Suspected gastroenteritis | >18y | bacterial vs viral | 108 | Interferon-γ | 67.0 | 63.0 | 0.65 |
| Ashkenazi-Hoffnung *et al.* (2018) | Inpatients and emergency department, Israel | Fever without source or respiratory infection | Any | bacterial vs viral | 314 | TNF-related apoptosis-inducing ligand+interferon gamma-induced protein-10+ C-reactive protein | 93.5 | 94.3 | |
| | | | | | | Interleukin-6 | 40.3 | 84.6 | |
| | | | | | | Lipocalin-2 | 71.1 | 77.5 | |
| Esposito *et al.* (2016b) | Inpatient unit, Italy | Children with community-acquired pneumonia | 4 m-14y | bacterial vs viral | 433 | Soluble triggering receptor expressed on myeloid cells-1 | 31.8 | 73.7 | 0.50 |
| | | | | | | Midregional proadrenomedullin | 78.0 | 35.7 | 0.58 |
| | | | | | | Midregional proatrial natriuretic peptide | 76.1 | 33.1 | 0.52 |
| Esposito *et al.* (2016a) | Intensive care unit, Italy | Radiologically confirmed community-acquired pneumonia | <14y | bacterial vs viral | 110 | Lipocalin-2 | 58.1 | 50.0 | 0.51 |
| | | | | | | Syndecan-4 | 31.1 | 86.1 | 0.54 |
| Liu *et al.* (2018) | Outpatient and inpatient units, 12 hospitals, China | Adult community-acquired pneumonia | >18y | bacterial vs viral | 124 | Platelet-derived growth factor BB | 82.1 | 70.7 | 0.70 |
| | | | | | | Interleukin-17A | 69.6 | 65.5 | 0.68 |
| | | | | | | Granulocyte colony-stimulating factor | 94.6 | 34.5 | 0.65 |
| | | | | | | Interleukin-10 | 42.9 | 82.8 | 0.63 |
| | | | | | | Bactericidal/permeability-increasing protein | 45.1 | 88.1 | 0.63 |
| | | | | | | Interleukin-2 | 85.5 | 48.0 | 0.62 |
| | | | | | | Basic fibroblast growth factor | 82.1 | 39.7 | 0.61 |

| | | | | | | | | | |
|---|---|---|---|---|---|---|---|---|---|
| Oved *et al.* (2015) | Emergency department, wards, Israel | Acute infection | Any | bacterial vs viral | 765 | TNF-related apoptosis-inducing ligand+interferon gamma-induced protein-10+ C-reactive protein | 87.0 | 90.0 | 0.94 |
| Qu *et al.* (2015) | Infectious disease department, China | Febrile patients | 18-85y | bacterial vs non-bacterial | 326 | Serum amyloid A | 42.0 | 58.0 | 0.68 |
| | | | | | | Interleukin-6 | 59.1 | 70.0 | 0.65 |
| Sanaei Dashti *et al.* (2017) | Inpatient unit, Iran | Suspected meningitis | 21days-144m | bacterial vs viral | 50 | Ferritin | 81.1 | 62.9 | 0.74 |
| Srugo *et al.* (2017) | Emergency department, Switzerland and Israel | Suspicion of acute infection | 3m-18y | bacterial vs viral | 361 | TNF-related apoptosis-inducing ligand+interferon gamma-induced protein-10+ C-reactive protein | 93.8 | 89.8 | |
| van der Does *et al.* (2018) | Emergency department, Netherlands | Fever | >18y | bacterial vs non-bacterial | 315 | TNF-related apoptosis-inducing ligand+interferon gamma-induced protein-10+ C-reactive protein | | | 0.73 |
| | | | | | | TNF-related apoptosis-inducing ligand+interferon gamma-induced protein-10+procalcitonin | | | 0.74 |
| | | | | | | TNF-related apoptosis-inducing ligand+interferon gamma-induced protein-10+ C-reactive protein+procalcitonin | | | 0.76 |
| | | | | | | TNF-related apoptosis-inducing ligand | 63.0 | 68.0 | 0.66 |
| | | | | | | Interferon gamma-induced protein-10 | 83.0 | 39.0 | 0.60 |
| van Houten *et al.* (2017) | Emergency department, Netherlands and Israel | Lower respiratory tract infection, fever without source | 2-60m | bacterial vs viral | 443 | TNF-related apoptosis-inducing ligand+interferon gamma-induced protein-10+ C-reactive protein | 86.7 | 91.1 | 0.90 |
| Wang *et al.* (2018) | Neurology department, China | Central nervous system infection | >18y | bacterial vs viral | 155 | s-100 protein | 80.0 | 85.0 | |
| Yanai *et al.* (2016) | General Medicine Department, Japan | Febrile patients | >18y | bacterial vs viral | 104 | Serum 2'-5'-oligoadenylate synthetase | | | 0.32 |
| Yang *et al.* (2018) | Medical Centre, China | Pneumonia in children | 1m-10 y | bacterial vs non-bacterial | 61 | Lectin microarray- haptoglobin-related protein | | | 0.72 |
| Yu *et al.* (2016) | Inpatient unit, Jilin university hospital, China | Signs and symptoms of acute infection | 10-71y | bacterial vs viral | 343 | Lipocalin-2- ELISA 1 (763/764 antibody) | 88.4 | 73.8 | 0.89 |
| | | | | | | Lipocalin-2- ELISA 2 (765/697antibody) | 98.5 | 94.2 | 0.97 |

| | | | | | | | | | |
|---|---|---|---|---|---|---|---|---|---|
| Zhou and Ye (2017) | Inpatient unit, Zhejiang university hospital, China | Pneumonia | <18y | bacterial vs viral | 515 | Interleukin-2 | 74.3 | 52.0 | 0.62 |
| | | | | | | Interleukin-4 | 18.3 | 100 | 0.60 |
| | | | | | | Interleukin-6 | 64.0 | 84.0 | 0.75 |
| | | | | | | Interleukin-10 | 88.6 | 56.0 | 0.24 |
| | | | | | | Tumour necrosis factor-alpha | 72.0 | 60.0 | 0.62 |
| | | | | | | Interferon-γ | 43.4 | 92.0 | 0.68 |
| | | | | | | Interleukin-6+interleukin-10 | 90.3 | 88.0 | 0.89 |
| Zhu *et al.* (2015) | Emergency department, China | Lower respiratory tract infection | 4-7y | bacterial vs viral | 96 | Interleukin-6 | | | 0.83 |
| Tan *et al.* (2023) | Emergency department, Netherlands | Fever within 72 hours | ≤18y | bacterial vs viral | 110 | TNF-related apoptosis-inducing ligand | | | 0.74 |
| | | | | | | Interferon gamma-induced protein-10 | | | 0.74 |
| | | | | | | Interferon-γ | | | 0.64 |
| | | | | | | Interleukin-4 | | | 0.60 |
| | | | | | | Lipocalin-2 | | | 0.72 |
| | | | | | | TNF-related apoptosis-inducing ligand+lipocalin-2 | | | 0.84 |
| | | | | | | TNF-related apoptosis-inducing ligand+lipocalin-2+interleukin-6 | | | 0.86 |
| Jackson *et al.* (2023) | Emergency departments, intensive care units, inpatient units, Europe | Fever within 72 hours | ≤18y | bacterial vs viral | 368 | E-selectin+ interleukin-18+ neural cell adhesion molecule-1+ lipocalin-2+ interferon-γ+ galectin-3 binding protein | 90.4 | 89·6 | 0.89 |
| Bartakova *et al.* (2019) | Infectious diseases department, Czech Republic | Signs of localised infection | >18y | bacterial vs viral | 116 | Calprotectin | | | 0.90 |
| | | | | | | Calgranulin C | | | 0.80 |
| Chen *et al.* (2020) | Inpatient unit, China | Pneumonia | 66 ± 20 y | bacterial vs viral | 108 | CD5 antigen like protein | 85.7 | 93.0 | 0.89 |
| Do *et al.* (2020) | Inpatient unit, National Hospital of paediatrics, Vietnam | Severe respiratory syncytial viral pneumonia | >1m <5y | viral vs co-infection | 81 | Interleukin-6 | 63.0 | 85.0 | 0.70 |
| Fang *et al.* (2020) | Inpatient unit, China | Signs of acute infection | ≤18y | bacterial vs viral | 574 | Lipocalin-2 | 67.2 | 83.1 | 0.81 |
| Havelka *et al.* (2020) | Infectious Disease Department, Sweden | Fever, symptoms of respiratory infection | >18y | bacterial vs viral | 279 | Calprotectin | 91.0 | 77.0 | 0.88 |
| | | | | | | Heparin binding protein | 49.0 | 78.0 | 0.63 |

| | | | | | | | | | |
|---|---|---|---|---|---|---|---|---|---|
| Imai *et al.* (2019) | Emergency department, Japan | Suspected systemic inflammatory response syndrome, bacteraemia | ≥70y | bacterial vs non-bacterial | 76 | Presepsin | 93.7 | 41.3 | 0.69 |
| Li, Yuan and Wang (2019a) | Inpatient unit, China | Hospitalised patients, with blood culture | 55.25 ± 21.75y | bacterial vs non-bacterial | 390 | Interferon-γ | 55.1 | 78.9 | 0.67 |
| | | | | | | Imterleukin-3 | 48.3 | 75.5 | 0.62 |
| | | | | | | Interferon-γ | 60.4 | 72.4 | 0.67 |
| | | | | | | Interleukin-17A | 45.8 | 78.9 | 0.65 |
| | | | | | | Macrophage inflammatory protein-1 beta | 66.7 | 75.6 | 0.70 |
| | | | | | | Tumour necrosis factor-alpha | 70.8 | 61.0 | 0.67 |
| | | | | | | Interleukin-3 | 69.1 | 73.0 | 0.72 |
| | | | | | | Interleukin-4 | 43.1 | 85.4 | 0.63 |
| Trouillet-Assant *et al.* (2020) | Paediatric emergency department, three hospitals, France | Febrile children | 7days-36m | bacterial vs viral | 101 | Interferon-alpha | | | 0.93 |
| | | | | | | Interferon-alpha+ C-reactive protein | | | 0.93 |
| Venge *et al.* (2019) | Infectious Disease Department, Sweden | Signs and symptoms of acute infections | >18y | bacterial vs viral | 288 | Lipocalin-2 | 63.0 | 75.0 | |
| | | | | | | Heparin binding protein | 57.0 | 63.0 | |
| | | | | | | Calprotectin | 68.0 | 65.0 | |
| | | | | | | Interferon gamma-induced protein-10 | 67.0 | 81.0 | |
| | | | | | | Thymidine kinase 1 | 38.0 | 79.0 | |
| | | | | | | TNF-related apoptosis-inducing ligand | 67.0 | 83.0 | |
| Dagys *et al.* (2022) | Paediatric emergency department, Lithuania | Fever <12 hours, signs in red column (NICE guidelines "Fever in under 5s") | <5y | bacterial vs viral | 70 | Interleukin-2 | 28.0 | 100 | 0.61 |
| | | | | | | Interleukin-6 | 68.0 | 69.0 | 0.70 |
| | | | | | | Soluble triggering receptor expressed on myeloid cell-1 | 54.0 | 86.0 | 0.65 |
| Nielsen *et al.* (2021) | Paediatric intensive care unit, United Kingdom | SBI present on admission | 0m-16y | bacterial vs non-bacterial | 657 | Lipocalin-2 | 61.0 | 75.0 | 0.68 |
| | | | | | | Resistin | 64.0 | 85.0 | 0.80 |
| Tasar *et al.* (2022) | Paediatrics department, Turkey | Pneumonia diagnosis | 1m-15y | bacterial vs viral | 41 | Endocan | 80.4 | 89.3 | 0.86 |

| | | | | | | | | | |
|---|---|---|---|---|---|---|---|---|---|
| Tsuchida *et al.* (2021) | Outpatient clinic and emergency department, Japan | Suspect bacterial infection | 62.1y-mean | bacterial vs non-bacterial | 1840 | Presepsin | 67.0 | 66.0 | 0.68 |
| Venge, Eriksson and Pauksen (2021) | Infectious diseases department or primary care unit, Sweden | Fever, signs and symptoms of acute respiratory infection | >18y | bacterial vs viral | 156 | Interferon gamma-induced protein-10 | | | 0.51 |
| | | | | | | Lipocalin-2 | | | 0.52 |
| | | | | | | TNF-related apoptosis-inducing ligand | | | 0.84 |
| | | | | | | Thymidine kinase 1 | | | 0.74 |
| | | | | | | Calprotectin | | | 0.87 |
| | | | | | | Heparin-binding protein | | | 0.53 |
| Chokkalla *et al.* (2023) | Emergency department, Texas | Acute febrile illness | ≤ 18y | bacterial vs viral | 60 | TNF-related apoptosis-inducing ligand+interferon gamma-induced protein-10+ C-reactive protein | 94.0 | 88.0 | |
| Halabi *et al.* (2023) | Emergency department, 3 hospitals, Israel | Fever, lower respiratory tract infection signs or symptoms for <7 days | >18y | bacterial vs viral | 415 | TNF-related apoptosis-inducing ligand+interferon gamma-induced protein-10+ C-reactive protein | 98.1 | 88.4 | |
| Lacroix *et al.* (2023) | Paediatric emergency department, Switzerland | Fever without source | <3y | bacterial vs viral | 241 | TNF-related apoptosis-inducing ligand+interferon gamma-induced protein-10+ C-reactive protein | | | 0.82 |
| | | | | | | Interferon gamma-induced protein-10 | | | 0.71 |
| Su *et al.* (2023) | Inpatient unit, China | Community-acquired pneumonia | <18y | bacterial vs non-bacterial (non-viral) | 263 | Serum Amyloid A | 78.3 | 93.1 | 0.88 |
| Wang *et al.* (2023) | Inpatient unit, Tongji hospital, China | Signs of infection | <18y | bacterial vs viral | 157 | Lipocalin-2 | 91.8 | 81.2 | 0.91 |

**Supplemental Table 2.** Summary of the Quality Assessment of Diagnostic Accuracy Studies-2 (QUADAS-2) assessment results of the included studies.

| | Risk of Bias | | | | Applicability Concerns | | |
|---|---|---|---|---|---|---|---|
| **Study** | Patient Selection | Index Test | Reference Standard | Flow and Timing | Patient Selection | Index Test | Reference Standard |
| Chalupa et al (2011)[30] | low | low | low | high | low | low | low |
| Conroy et al (2014)[31] | high | low | high | high | high | low | high |
| Elsing et al (2011)[32] | low | low | low | low | low | low | low |
| Haran et al (2013)[33] | low | low | high | low | low | low | high |
| Katoh et al (2014)[34] | unclear | low | low | high | unclear | low | low |
| Mansour et al (2011)[35] | low | low | low | low | low | low | low |
| Siahanidou et al (2014)[36] | low | low | low | low | low | low | low |
| Te Witt et al (2012)[37] | unclear | low | high | high | high | low | high |
| Ten Oever et al (2012)[38] | low | low | low | low | low | low | low |
| Weh et al (2013)[39] | low | low | low | low | low | low | low |
| Ashkenazi-Hoffnung et al (2018)[40] | low | low | low | low | low | low | low |
| Esposito et al (2016)[41] | low | low | low | low | low | low | low |
| Esposito et al (2016)[42] | low | low | low | low | low | low | low |
| Liu et al (2018)[43] | low | low | low | low | low | low | low |
| Oved et al (2015)[22] | low | low | low | low | low | low | low |
| Qu et al (2015)[44] | low | low | high | high | low | low | high |
| Sanaei et al (2017)[45] | low | low | low | low | low | low | low |
| Srugo et al (2017)[46] | low | low | low | low | low | low | low |
| van der Does et al (2018)[47] | high | low | high | low | low | low | high |
| van Houten et al (2017)[48] | low | low | low | low | low | low | low |
| Wang et al (2018)[49] | low | low | low | low | low | low | low |
| Yanai et al (2016)[50] | low | low | low | low | low | low | low |
| Yang et al (2018)[51] | unclear | low | low | high | low | low | low |
| Yu et al (2016)[52] | low | low | low | low | low | low | low |
| Zhou et al (2017)[53] | low | low | low | high | low | low | low |
| Zhu et al (2015)[54] | low | low | low | high | low | low | low |
| Tan et al (2023)[18] | high | low | low | high | low | low | low |
| Jackson et al (2023)[55] | low | low | low | low | low | low | low |
| Bartakova et al (2019)[56] | low | low | low | high | low | low | low |
| Chen et al (2020)[57] | high | high | high | high | low | low | unclear |
| Do et al (2020)[58] | low | low | low | high | high | low | low |
| Fang et al (2020)[59] | low | low | low | low | low | low | low |
| Havelka et al (2020)[60] | low | low | low | high | low | low | low |
| Imai et al (2019)[61] | low | low | low | low | low | low | low |
| Li et al (2019)[62] | low | low | low | low | low | low | low |
| Trouillet-Assant et al (2020)[63] | low | low | low | low | low | low | low |
| Venge et al (2019)[64] | low | low | low | low | low | low | low |
| Dagys et al (2022)[65] | low | low | low | low | low | low | low |
| Nielsen et al (2021)[66] | low | low | low | low | low | low | low |
| Tasar et al (2022)[67] | high | low | high | high | low | low | high |
| Tsuchida et al (2021)[68] | low | low | low | high | low | low | low |
| Venge et al (2021)[69] | low | low | low | high | low | low | low |
| Chokkalla et al (2023)[70] | low | low | low | low | low | low | low |
| Halabi et al (2023)[71] | low | low | low | low | low | low | low |
| Lacroix et al (2023)[72] | low | low | low | low | low | low | low |
| Su et al (2023)[73] | low | low | low | high | low | low | low |
| Wang et al (2023)[74] | low | low | low | low | low | low | low |
